# Quantification of Protein Biomarkers in SonoPODography Fluid Using Single Molecule Arrays for Endometriosis

**DOI:** 10.1101/2025.09.19.25336201

**Authors:** Kezia E. Suryoraharjo, Shay M. Freger, Sophia M. Alonzi, Anthony Makwanda, Mathew Leonardi, Alana F. Ogata

## Abstract

**BACKGROUND:** Patients with endometriosis experience chronic, inflammatory symptoms including severe pelvic pain and infertility, caused by endometrial-like tissue growth outside the uterus. Endometriosis remains poorly understood, largely attributed to disease heterogeneity and dependence on qualitative technologies for research. There is an unmet clinical need for quantitative, biomarker-based methods for improved endometriosis diagnosis, prognosis, and treatment. We describe the biomarker analysis of a novel biofluid for endometriosis research, sonoPODography (SPG) fluid, which is a fluid collected using culdocentesis from the rectouterine pouch upon saline infusion through the uterus and fallopian tubes.

**METHODS:** Single molecule array (Simoa) assays were used to quantify TNFα, IL-1β, VEGF, and CA125 in SPG fluid of 33 endometriosis patients. Simoa assays were validated using dilution-linearity and spike-and-recovery experiments in pooled and individual SPG fluid samples from endometriosis patients. The agreement of CA125 measurements by Simoa and enzyme-linked immunosorbent assay (ELISA) was assessed using Spearman correlation and Bland-Altman analyses.

**RESULTS:** Simoa assays were validated in SPG fluid and dilution factors 2x (TNFα), 4x (IL-1β), 8x (VEGF), 600x (CA125) were selected for biomarker quantification. Median biomarker concentrations in SPG fluid from endometriosis patients were 0.276 pg/mL (TNFα), 0.21 pg/mL (IL-1β), 30.55 pg/mL (VEGF), and 1,810 pg/mL (CA125). Simoa and ELISA CA125 measurements were highly correlated (ρ = 0.80, *p* < 0.0001).

**CONCLUSION:** We present the feasibility and potential of SPG fluid as a novel source of biomarkers for endometriosis, which can enable quantitative clinical technologies for endometriosis diagnosis, prognosis, and treatment.

## Introduction

Endometriosis is a chronic, inflammatory disease where endometrial-like tissue grows outside the uterus. Patients with endometriosis commonly experience severe pelvic pain, infertility, and other debilitating health problems (1). The World Health Organization estimates that endometriosis affects 10% of people assigned female at birth globally, with a diagnostic delay of 6 years (2) that translates to over 2 million disability-adjusted life years (DALY) in 2019 (3, 4). Historically, the gold standard for diagnosing and studying endometriosis is laparoscopic surgery followed by histopathological confirmation (5), which is invasive, costly, and dependent on surgeon expertise in identifying and excising lesions (6). Recognizing the drawbacks from exclusive reliance on laparoscopic surgery, guidelines for endometriosis such as those by the European Society of Human Reproduction and Embryology (ESHRE) in 2022, the Society of Obstetricians and Gynaecologists Canada (SOGC) in 2022, and the American College of Obstetricians and Gynecologists (ACOG) in 2010 have recommended imaging methods, such as ultrasonography (US) and magnetic resonance imaging (MRI), and empirical treatment as alternative options to diagnose endometriosis (7–9). Despite advancements in imaging methods, the diagnosis and stratified treatment of endometriosis remain a challenge due to the heterogeneous nature of the disease, which shows varying presentation of symptoms and lesions (1, 10, 11). Additionally, there remains a poor understanding of endometriosis pathophysiology due to the presence of multiple common comorbidities and the lack of non-invasive, quantitative tools.

Biomarker-based methods for endometriosis have been of significant interest to support etiology research and enhance the diagnosis, prognosis, and treatment of endometriosis. Biomarker discovery efforts for endometriosis include the analysis of plasma (12, 13), serum (12, 14–18), and peritoneal fluid (14, 15, 18–21), however no biomarkers have shown sufficient clinical sensitivity and specificity for diagnostics. In comparison to blood, peritoneal fluid is a promising source for endometriosis-specific biomarkers as it reflects the local pelvic environment where endometriotic lesions are typically present (22). Biomarkers such as tumor necrosis factor-alpha (TNFα) (15), interleukin-6 (IL-6) (15, 19, 23), monocyte chemotactic protein-1 (MCP-1) (19), and cancer antigen 125 (CA125) (14) have been reported to be elevated in the peritoneal fluid of patients with endometriosis compared to those without endometriosis. However, peritoneal fluid collection is primarily performed via laparoscopic surgery (24), and its invasive nature poses challenges for routine diagnostic testing. Alternatively, peritoneal fluid collected by a less invasive method, such as culdocentesis, often yields insufficient volume for biomarker analysis (21, 24, 25).

Leonardi *et al.* introduced a novel technique to visualize endometriotic lesions in the pelvic posterior compartment via transvaginal US (TVUS) termed saline-infusion sonoPODography (SPG) (26, 27). SPG involves the introduction of saline into the rectouterine pouch through the uterus and fallopian tubes, which supplements acoustic enhancement provided by naturally present peritoneal fluid and enhances identification of endometriotic lesions for cases where peritoneal fluid volume is insufficient for visualization. The SPG method results in a saline-infused peritoneal fluid, termed here as SPG fluid, that is in direct contact with endometriotic lesions and can be collected non-surgically by TVUS-guided culdocentesis (TVS-C). SPG fluid offers a novel, less invasive and therefore accessible sample for endometriosis biomarker research.

This study describes the analysis of SPG fluid for endometriosis biomarker quantification using single molecule arrays (Simoa). Simoa is an ultrasensitive technology for the detection of analytes in biofluids, also known as the digital enzyme-linked immunosorbent assay (ELISA), that provides over 100x higher sensitivity than traditional ELISA (28) and has been demonstrated for biomarker analysis in blood (29), cerebrospinal fluid (30), and saliva (31). Simoa achieves sub-femtomolar (10^-15^ M) limits of detection with a dynamic range over four orders of magnitude that can enable robust assessment of biomarkers in novel biofluids such as SPG fluid (32). Simoa utilizes a sandwich immunoassay format composed of two antibodies that target a single analyte. Each immunocomplex (microbeads, capture antibody, analyte, detection antibody, streptavidin-β-galactosidase) is physically isolated into femtoliter-sized wells, enabling the digital counting of single molecules. Here, we describe SPG fluid collection, sample preparation, and validation of Simoa assays in SPG fluid for the quantification of TNFα, IL-1β, vascular endothelial growth factor (VEGF), and CA125 in SPG fluid. Through this work, we provide foundational insights into the utility of SPG fluid as a novel source for endometriosis biomarker research.

## Materials and Methods

### Study Participants

Participants were prospectively recruited from McMaster University Medical Centre, a tertiary, university-affiliated gynecologic surgical center with clinical expertise in endometriosis, chronic pelvic pain, and general gynecologic concerns. Premenopausal individuals aged 18–55 years who were scheduled to undergo laparoscopic surgery for any benign indication were eligible for inclusion. Surgical indications included, but were not limited to, suspected endometriosis with planned excision, hysterectomy (with or without salpingo-oophorectomy), myomectomy, and fertility-related procedures. Exclusion criteria comprised known or suspected malignancy, current pregnancy, and any contraindications to SPG and/or TVS-C. These included an obliterated rectouterine pouch, history of hysterectomy or bilateral salpingectomy, non-patent fallopian tubes (as assessed by preoperative imaging or surgical history), or extensive anatomic distortion unrelated to endometriosis (e.g., multifibroid uterus with obliteration of normal pelvic landmarks). Additional exclusion criteria were intraoperative findings precluding safe instillation or aspiration of the posterior rectouterine pouch, such as dense filmy adhesions or an inaccessible vaginal fornix.

Participants with endometriosis were identified as those undergoing excision of suspected disease based on intraoperative surgical findings. In the participants with endometriosis, SPG and TVS-C were performed sequentially following induction of anesthesia and prior to any tissue excision. Demographic and clinical information were collected at the time of enrollment using standardized case report forms and validated patient-reported questionnaires. Collected data included age, body mass index (BMI), menstrual history, pelvic pain symptoms, use of hormonal therapies, and history of prior surgical diagnosis of endometriosis. This study was approved by the institutional review board of each participating institution (UTM#44938/2023-04-10/2026-12-31, HiREB#14503/2023-03-10/2026-03-20) and adhered to the principles of the Declaration of Helsinki.

### SPG Fluid Patient Sample Collection

At the time of scheduled laparoscopic surgery, participants underwent SPG immediately following induction of general anesthesia. SPG was performed as previously described by Leonardi *et al*., wherein sterile saline was transvaginally instilled into the posterior rectouterine pouch under continuous TVUS guidance to distend the rectouterine pouch and enhance real-time visualization (Supplemental Information, Methods) (26, 27).

Intraoperatively, endometriosis was characterized visually by the attending surgeon and categorized as superficial endometriosis (SE), deep endometriosis (DE), ovarian endometrioma (OE), or a combination thereof. This classification was based on anatomic location, depth of infiltration, and lesion morphology, consistent with international surgical definitions. Where feasible, lesions were excised and submitted for histopathologic confirmation. Cases were considered definitive for endometriosis only if the disease was surgically visualized and confirmed on histology.

### Sample Processing

All SPG fluid samples were processed within one hour of collection, where samples were immediately centrifuged at 3,000 rpm for 20 minutes at room temperature to separate the cellular pellet from the acellular supernatant. The supernatant was aspirated and aliquoted into sterile, low-retention 0.5 mL cryovials, and immediately stored at −80°C for subsequent biomarker analyses.

### Simoa Assays

All biomarker measurements in this study were conducted using single molecule array (Simoa) assays on the HD-X Analyzer (Quanterix) according to the manufacturer’s instructions (Supplemental Information, Methods). All calibrators and samples were run in duplicates. TNFα levels were quantified using the Simoa TNFα Advantage Kit (cat# 101580, Quanterix), IL-1β levels were quantified using the Simoa IL-1β 3.0 Kit (cat#103464, Quanterix). VEGF and CA125 levels were quantified using Simoa assays developed in-house (Supplemental Information, Methods). For the VEGF and CA125 Simoa assays, capture antibody-conjugated beads were diluted to a final concentration of 2 x 10^7^ beads/mL in Bead Diluent (Quanterix). For the VEGF assay, biotinylated detector antibodies were diluted to 0.3 µg/mL in Homebrew Detector/Sample Diluent (Quanterix) and Streptavidin-β-galactosidase (SβG) Concentrate (Quanterix) was diluted to 75 pM in SβG Diluent (Quanterix). For the CA125 assay, biotinylated detector antibodies were diluted to 0.1 µg/mL and SβG Concentrate was diluted to 150 pM in SβG Diluent. Calibrators were prepared by performing serial dilutions of recombinant protein standards (VEGF cat# BT-VEGF, R&D Systems; CA125 cat# DY5609-05, R&D Systems) in Homebrew Detector/Sample Diluent, along with high and low controls. SPG fluid samples were diluted in Homebrew Detector/Sample Diluent. Both VEGF and CA125 were measured with three-step assays. 25 µL of the bead solution and 100 µL of sample were incubated for 15 minutes, followed by a wash step with System Wash Buffer 1 (Quanterix). The beads were resuspended in 100 µL of biotinylated detector antibodies and incubated for 5 minutes 15 seconds, followed by a wash step with System Wash Buffer 1. The beads were then resuspended in 100 µL of SβG and incubated for 5 minutes 15 seconds, followed by six wash steps with System Wash Buffer 1 and a final wash step with System Wash Buffer 2 (Quanterix). The beads were resuspended in either 50 µL of Resorufin-β-D-galactopyranoside (RGP) Reagent for the VEGF assay or 25 µL of RGP Reagent for the CA125 assay before being loaded into the Simoa disc (Quanterix). The microwell array was subsequently sealed with Simoa Sealing Oil (Quanterix) and subjected to image analysis. Average Enzyme per Bead (AEB) values were fit against biomarker concentrations using a 4-parameter logistic curve weighted by 1/y^2^ to generate calibration curves. For all Simoa assays, two quality control samples were measured in each independent run with errors below 20%. Concentration determination of the samples was done automatically by the software on the HD-X Analyzer utilizing the calibration curves.

### Simoa Assay Validation

Frozen SPG samples (−80°C) were thawed on ice. Samples from five patients were combined to constitute the pooled samples. The samples were centrifuged at 3,000 x g for 15 minutes at 4°C, and the supernatant was collected. For dilution linearity, recombinant protein of each biomarker was spiked into the pooled sample, followed by Simoa measurements in samples serially diluted with Homebrew Detector/Sample Diluent.

Spike-and-recovery experiments were conducted on pooled samples as well as three individual samples. Percent recoveries were examined in the samples with spiked recombinant protein. For each biomarker, three concentrations were chosen at low, mid, and high-range concentrations at each dilution factor selected. The percent recoveries were calculated by subtracting the mean measured concentration of the samples without the spiked protein from the mean measured concentration of the spiked samples, then dividing by the concentration of recombinant protein spiked into the samples and multiplying by 100%.

### Simoa Biomarker Quantification

Frozen SPG samples (−80°C) were thawed on ice. The samples were centrifuged at 3,000 x g for 15 minutes at 4°C, and the supernatant was collected. For quantification using Simoa assays, samples were diluted 2x (TNFα), 4x (IL-1β), 8x (VEGF), and 600x (CA125) in Homebrew Detector/Sample Diluent. Biomarker concentrations of samples were determined using calibration curves generated on the same day of quantification.

### Data Analysis

Data analysis was performed in GraphPad Prism version 10.4.2. All figures were generated using GraphPad Prism version 10.4.2. For all the data presented in this study otherwise noted, duplicate measurements per sample were obtained and the mean values were plotted. Error bars represent the standard deviation of the measurements. The limit of detection (LOD) was calculated as the concentration corresponding to three standard deviations (SD) of the first calibrator added to the background signal. The threshold for quantification was set to be cumulative LOD + 3SD.

The Shapiro-Wilk test was used to evaluate the Gaussian distribution of the quantification data. The Mann-Whitney test was used for the quantification data with non-normal distribution. The correlation coefficient of CA125 concentrations measured by Simoa and ELISA was calculated using the Spearman correlation analysis. The difference of CA125 measurements with the two methods was determined by Bland-Altman analysis. Statistically significant differences were considered when the two-tailed *p*-value was < 0.05.

SPG fluid samples were collected via TVUS-guided culdocentesis (Figure 1a), as described in the methods, and resulted in 33 participants classified as endometriosis cases based on laparoscopic surgery and histopathologic confirmation. Demographic and clinical data can be found in Table 1. SPG fluid samples were processed for protein biomarker quantification using Simoa assays. We selected the biomarkers TNFα, IL-1β, VEGF, and CA125 for analysis, which were previously reported to show diagnostic potential in peritoneal fluid from endometriosis patients. The cytokines TNFα and IL-1β are implicated in endometriosis development due to their promotion of cell proliferation, adhesion, and angiogenesis (15). VEGF is an angiogenesis marker involved in the development of blood vessels (33). CA125 is a common marker for gynecological cancers and proposed to be related with severity of pelvic adhesions (34). Simoa assay performance for TNFα, IL-1β, VEGF, and CA125 was assessed on *N* independent days (Supplemental Tables 1-4) based on calibration curves and LOD, defined as the mean signal of the blank plus three times the standard deviation of the first calibrator. The average LOD was 0.010 ± 0.008 pg/mL (*N* = 9) for the TNFα Simoa assay, 0.01 ± 0.01 pg/mL (*N* = 9) for the IL-1β Simoa assay, 1 ± 1 pg/mL (*N* = 12) for the VEGF Simoa assay, and 0.3 ± 0.3 pg/mL (*N* = 15) for the CA125 Simoa assay (Figures 1b-1e).

**Figure 1.**
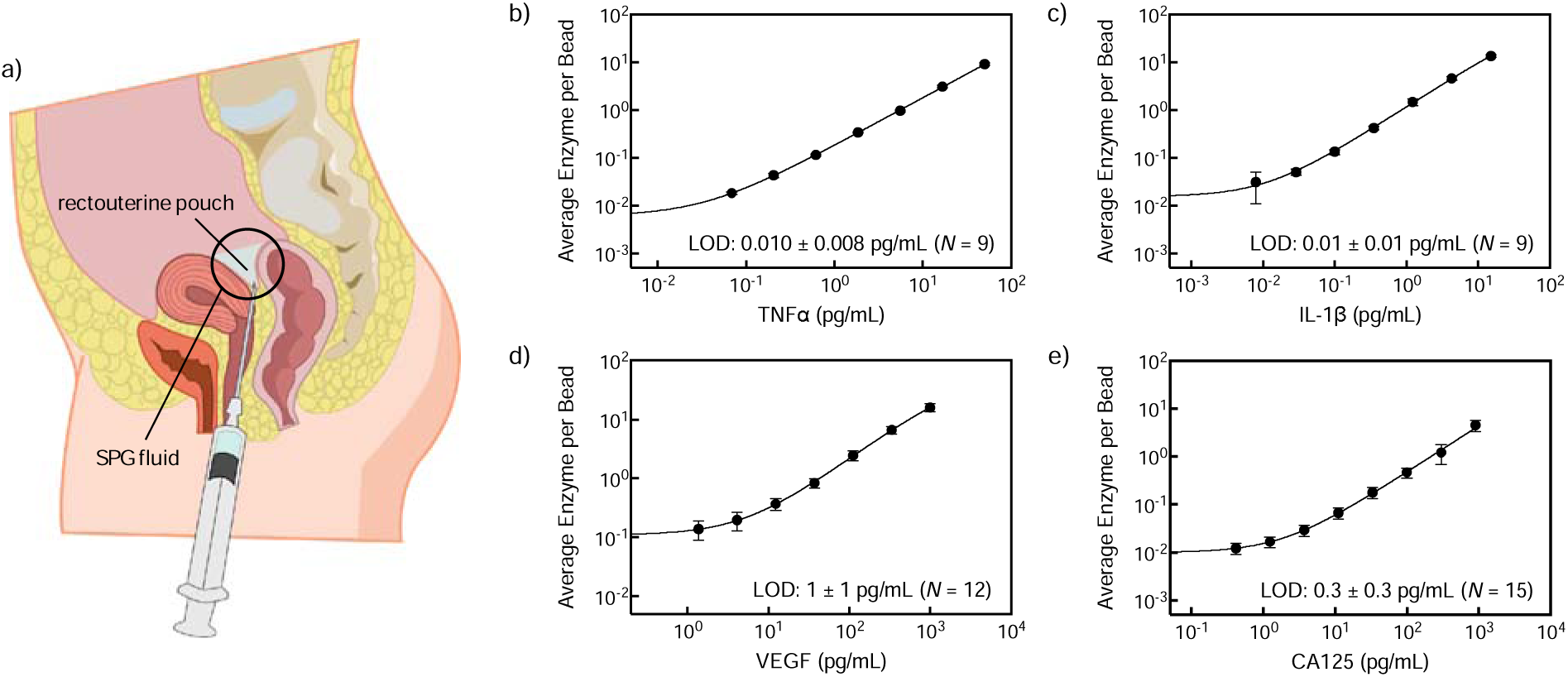
a) Sagittal diagram of the female pelvic cavity. SPG fluid is saline infused to the rectouterine pouch, collected via culdocentesis. b-e) The plotted calibration curves represent *N* independently generated Simoa calibration curves: b) TNFα (*N* = 9), c) IL-1β (*N* = 9), d) VEGF (*N* = 12), e) CA125 (*N* = 15). Error bars represent the standard deviation at each concentration across *N* replicates. LODs of each assay inset on each plot represent the mean of *N* independent LODs.

**Table 1.** Demographic characteristics of study participants.

| Characteristic | Patients with Endometriosis<br>($N = 33$) |
| --- | --- |
| Age (years), mean ± SD | $36 \pm 9$ |
| Sex |  |
| Assigned female at birth, n (%) | 33 (100%) |
| Weight (kg), mean ± SD | $73 \pm 20$ |
| Height (m), mean ± SD | $1.66 \pm 0.08$ |
| Body-mass index ($\text{kg}/\text{m}^2$), mean ± SD | $27 \pm 7$ |
| Gravidity, median (IQR) | 0 (0-2) |
| Parity, median (IQR) | 0 (0-1) |
| Saline volume introduced (mL), mean ± SD | $79 \pm 4$ |
| Estimated initial peritoneal fluid (mL), mean ± SD | $3 \pm 5$ |

TNFα, IL-1β, VEGF, and CA125 Simoa assays were then validated in pooled SPG fluid from endometriosis patients through dilution linearity and spike-and-recovery experiments. For dilution linearity, recombinant proteins specific to each assay were added to pooled SPG fluid samples followed by dilution and measurement of response (Figure 2). The linear dilution range was identified as 1x to 32x with R^2^ > 0.99 for the TNFα Simoa assay, 1x to 32x with R^2^ = 0.98 for the IL-1β Simoa assay, 4x to 32x with R^2^ = 0.98 for the VEGF Simoa assay, and 100x to 1000x with R^2^ > 0.99 for the CA125 Simoa assay.

**Figure 2.**
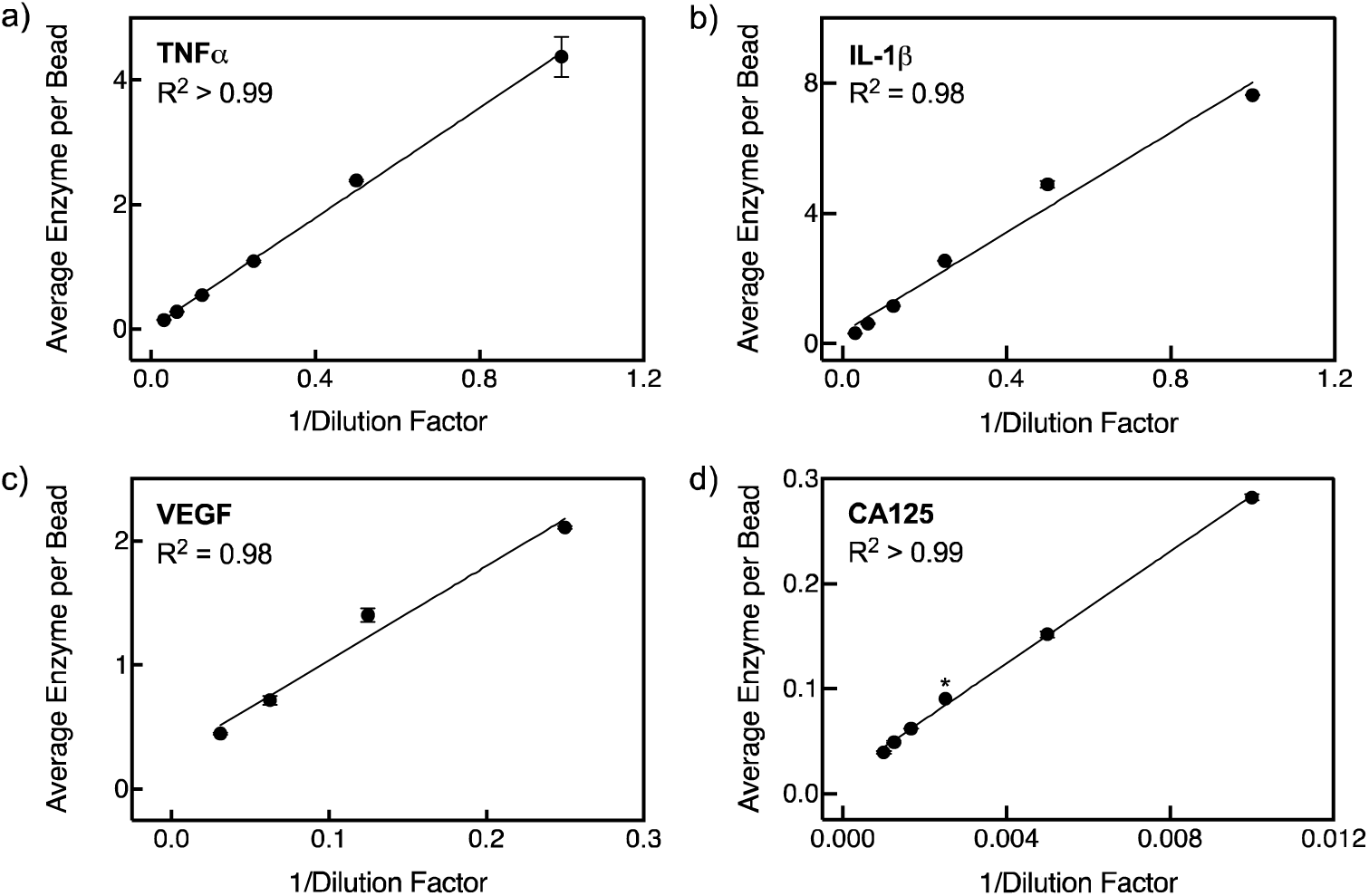
Linear response of four Simoa assays in SPG fluid. a) TNFα assay response over 1x to 32x dilution factors. b) IL-1β assay response over 1x to 32x dilution factors. c) VEGF assay response over 4x to 32x dilution factors. d) CA125 assay response over 100x to 1000x dilution factors. Data are the average of two replicates with error bars representing the standard deviation. Point marked with (*) indicates one replicate.

We then carried out spike-and-recovery experiments using SPG fluid spiked with low, mid, and high concentrations of recombinant proteins to determine the optimal dilution factor for Simoa assays in SPG fluid that minimized matrix effects with the least dilution possible. Three dilution factors in the linear dilution range were tested for spike-and-recovery rates in pooled SPG fluid from endometriosis patients (Figures 3a-d). Based on the dilution factor that resulted in a recovery of 80 – 120% in pooled samples, spike-and-recovery experiments were repeated in three individual patient SPG samples (Figures 3e-h). Optimal dilution factors were identified as 2x for TNFα, 4x for IL-1β, 8x for VEGF, and 600x for CA125. Percent recoveries for individual patient samples were within 80-120% for all assays with the exception of the low spike concentrations for two of the three patient samples for TNFα, IL-1β, and VEGF. The variability in percent recoveries is likely due to individual differences in SPG fluid matrix composition, with effects being more pronounced at low spike concentrations (0.2 pg/mL TNFα, 0.05 pg/mL IL-1β, 3 pg/mL VEGF, 1 pg/mL CA125). While further dilution can improve recoveries, we proceeded with dilution factors 2x for TNFα, 4x for IL-1β, 8x for VEGF, and 600x for CA125 as they provided optimal compromise between minimizing matrix effects and circumventing dilution the biomarkers to undetectable concentrations. To enable signal differentiation from noise and robust protein biomarker quantification, we established a stringent detection threshold to be the average LOD across *N* independently generated calibration curves of each Simoa assay plus three standard deviations of the LOD values across *N* curves (LOD + 3SD).

**Figure 3.**
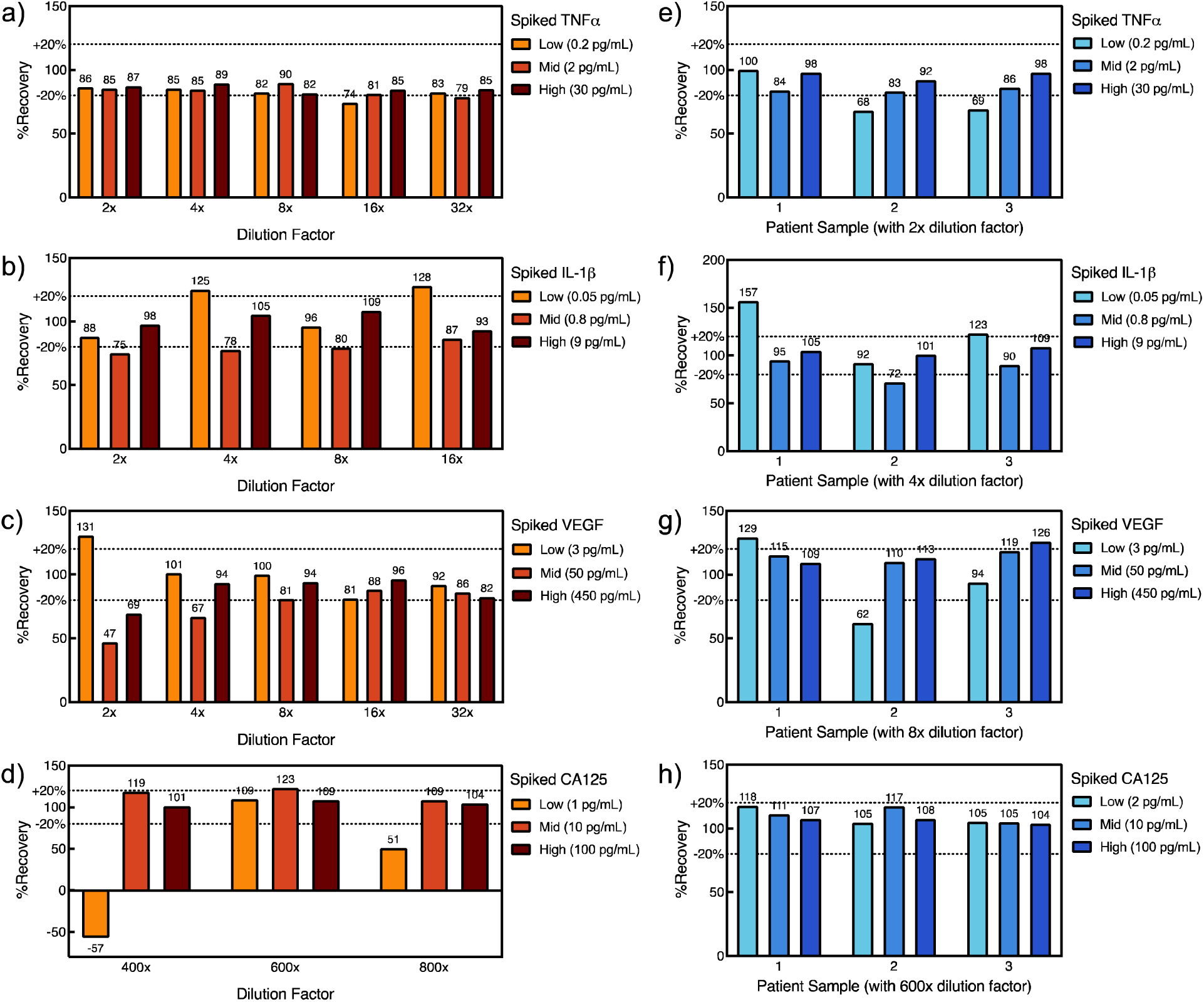
Percent recoveries of four Simoa assays in pooled (a-d) and three individual (e-h) patient SPG fluid samples. a, e) Percent recoveries of TNFα assay. b, f) Percent recoveries of IL-1β assay. c, g) Percent recoveries of VEGF assay. d, h) Percent recoveries of CA125 assay.

Validated TNFα, IL-1β, VEGF, and CA125 Simoa assays were then used to analyze biomarker levels individual SPG fluid samples from 33 endometriosis patient. Using LOD + 3SD as the cut-off value, TNFα was detected in 26 samples (79%), IL-1β was detected in 9 samples (27%), VEGF was detected in 13 samples (39%), CA125 was detected in 30 samples (91%) (Figure 4).

**Figure 4.**
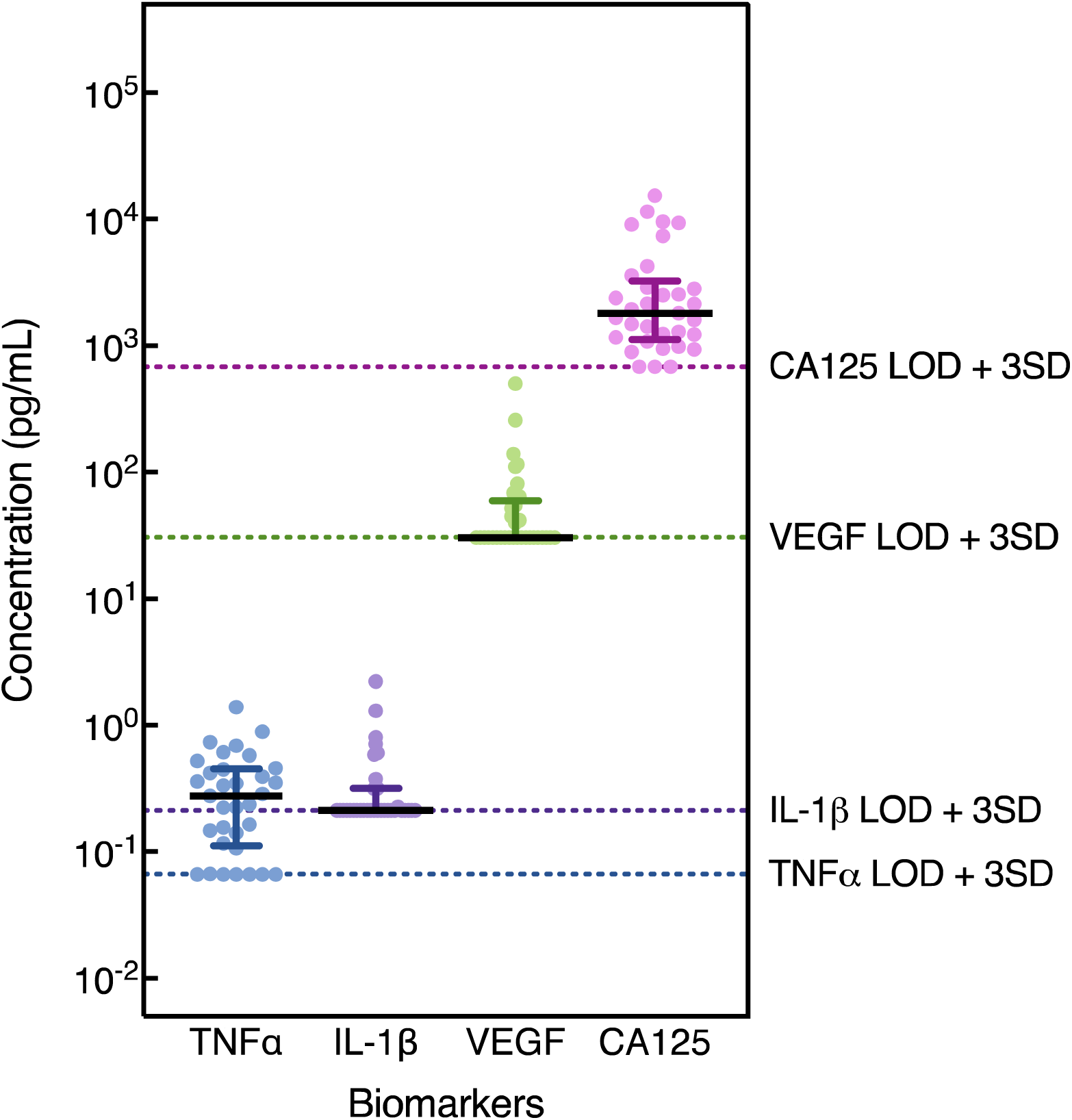
Quantification of TNFα, IL-1β, VEGF, and CA125 in individual endometriosis patient SPG fluid samples (*N* = 33). Data are the average of two replicates, corrected by dilution factors. Black lines indicate medians, error bars indicate interquartile range (IQR). Coloured dotted lines indicate LOD + 3SD corrected by dilution factors.

The median TNFα concentration was 0.276 pg/mL with IQR 0.112-0.453 pg/mL, which corroborates previously reported TNFα concentrations in peritoneal fluid with median (IQR) levels of 54.83 (26.25-63.46) pg/mL in endometriosis cases and 0.00 (0.00-9.61) pg/mL in non-endometriosis cases (15). The median IL-1β concentration at 0.21 pg/mL with IQR 0.21-0.32 pg/mL also matched previously reported concentrations in peritoneal fluid with median (IQR) levels of 4.30 (0.00-13.67) pg/mL in endometriosis cases and 3.53 (0.00-35.96) pg/mL in non-endometriosis cases (15). The median VEGF concentration was 30.55 pg/mL with IQR 30.55-59.54 pg/mL. The range of VEGF concentrations were lower by an order of magnitude compared to VEGF concentrations in peritoneal fluid that have been reported with mean levels of 24.05 ± 15 ng/mL in endometriosis cases and 13.25 ± 7.2 ng/mL in non-endometriosis cases (20). The median CA125 concentration was 1,810 pg/mL, with an IQR 1,126-3,241 pg/mL. Compared to the cytokines TNFα and IL-1β, CA125 levels in SPG fluid were over three orders of magnitude higher, likely reflecting its greater molecular weight. As CA125 is secreted by cells lining the endometrium, fallopian tubes, and peritoneal surfaces (35), saline infused to the rectouterine pouch through these structures may sample high levels of CA125, contributing to the elevated concentrations of CA125 in SPG fluid measured in this study.

Quantification of biomarkers by Simoa showed that TNFα, IL-1β, and VEGF concentrations were lower than the standard detection limits of ELISA (36–38); however, CA125 levels were present at concentrations within ELISA detection limits and enabled the assessment of agreement of the CA125 Simoa assay and a CA125 ELISA. For the CA125 ELISA assay, we validated the assay in SPG fluid (Supplemental Information, Methods) and selected an 8x dilution factor based on dilution linearity and spike-and-recovery experiments in SPG fluid (Supplemental Figure 1). The median values of CA125 levels measured by the Simoa and ELISA were not significantly different (Figure 5a) and CA125 measurements across the two methods showed strong correlation (ρ = 0.80, *p* < 0.0001, Figure 5b). Bland-Altman analysis revealed a systematic bias of −854.7 pg/mL for Simoa compared to ELISA, with discrepancy between assay measurements being more pronounced at concentrations above 3,000 pg/mL (Figure 5b-c). We attribute this bias to the two-order-of-magnitude difference in dilution factors between the validated Simoa (600x) and ELISA (8x) CA125 assays in SPG fluid. These differing dilutions may account for the assays’ varying susceptibility to matrix effects such as cross-reactivity and non-specific binding.

**Figure 5.**
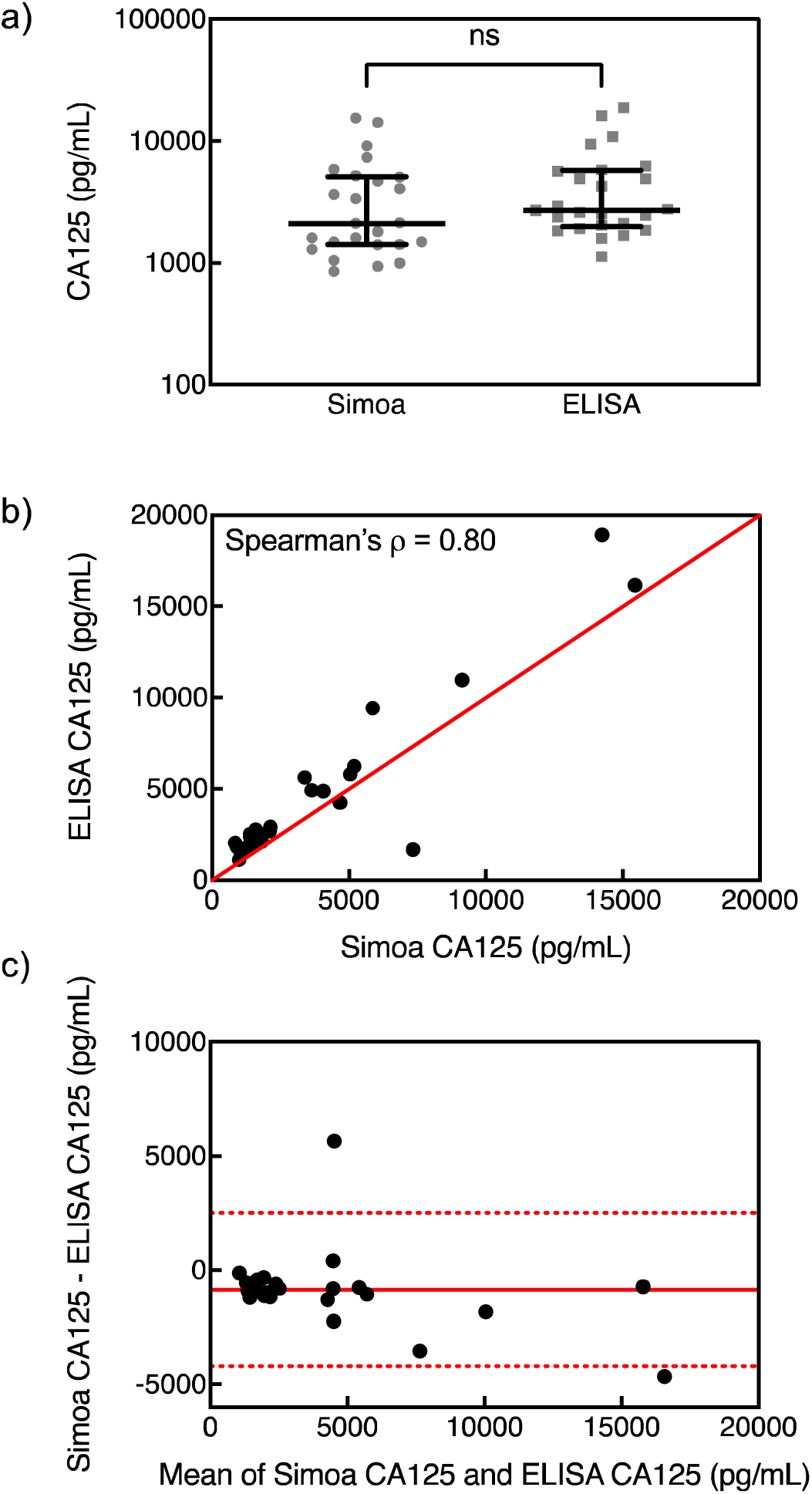
Comparison of CA125 levels measured by Simoa and ELISA. a) CA125 concentrations measured by Simoa and ELISA. Lines indicate median, error bars indicate IQR. Difference in median was assessed by the Mann-Whitney test. b) CA125 concentrations measured by ELISA against Simoa. Red line indicates line of identity. c) Bland-Altman plot for CA125 levels measured by Simoa versus ELISA. Solid red line indicates bias, dotted red lines indicate 95% CI. Data are the average of two replicates.

## Conclusion

Research in endometriosis can be accelerated by the advent of quantitative technologies to measure biomarkers for diagnostics, prognostics, and stratified treatment. In this study, we presented the collection and biomarker analysis of a novel SPG fluid for endometriosis research. We validated TNFα, IL-1β, VEGF, and CA125 Simoa assays in SPG fluid and quantified biomarker levels in SPG fluid samples from 33 endometriosis patients. For each biomarker in SPG fluid, concentrations varied up to two orders of magnitude across individuals, highlighting the potential to quantify the dynamics of biomarker concentrations amongst patient groups and for longitudinal monitoring. A critical next step for this research includes biomarker quantification in SPG samples from a sample size with sufficient power for diagnostic accuracy studies composed of cases and controls, cases by subtype of disease, or cases classified by symptom presentation. In this regard, an advantage of SPG fluid for biomarker analysis is that collection of SPG fluid is an accessible procedure in outpatient facilities, enabling the high-throughput collection of control case samples from non-endometriosis patients. Secondly, SPG is a cutting-edge method to accurately detect the SE subtype, which is a currently understudied subtype due to challenges in diagnosis by gold standard methods. The integration of SPG and SPG fluid biomarker analyses can expand research on the SE subtype, which is prevalent in 80% of endometriosis cases. We preliminarily demonstrate the feasibility of performing biomarker analysis in SPG fluid using Simoa for applications in diagnostics (Supplemental Table 5, Supplemental Figures 2-3), disease stratification (Supplemental Figure 4), and symptom presentation (Supplemental Figures 5-9). Future work must also consider biological variables and confounding factors that may influence biomarker concentrations. For instance, the menstrual cycle has been reported to influence levels of IL-12(p70), intercellular adhesion molecule-1 (ICAM-1), growth-related oncogene-alpha (GRO-α), and eotaxin in peritoneal fluid (19). Moreover, as with other biofluid-based analysis such as those in blood and urine, variation in dilution can result in measurement bias. In SPG, volumes of saline introduced range 79 ± 4 mL and does not account for the two-orders-in-magnitude range of biomarker concentrations in SPG measured in this study. However, the variation in saline volume combined with differences of pre-existing peritoneal fluid volume may hinder the realization of sufficient clinical sensitivity and specificity for clinical applications, necessitating the future optimization and evaluation of standardization protocols of SPG fluid biomarker analyses. Future work will be critical in advancing assay development and biomarker validation in SPG fluid, contributing to the goal of developing quantitative technologies for endometriosis diagnosis, prognosis, and treatment. This work provides the first step towards that goal by providing the technical foundation for the analysis of SPG fluid for biomarker-based methods that can open avenues in endometriosis research.

## Author Contributions

Suryoraharjo, Kezia Erina (Data curation (Equal), Formal analysis (Lead), Investigation (Equal), Validation (Equal), Visualization (Lead), Writing - original draft (Equal), Writing - review & editing (Equal)). Freger, Shay Michael (Data curation (Equal), Investigation (Equal), Writing - original draft (Supporting), Writing - review & editing (Supporting)). Alonzi, Sophia Maria (Investigation (Equal), Validation (Supporting), Writing - review & editing (Supporting)). Makwanda, Anthony (Investigation (Supporting), Validation (Supporting)). Leonardi, Mathew (Conceptualization (Equal), Funding acquisition (Equal), Investigation (Equal), Methodology (Lead), Project administration (Equal), Resources (Equal), Writing - review & editing (Supporting)). Ogata, Alana (Conceptualization (Equal), Funding acquisition (Equal) Project administration (Equal), Resources (Equal), Supervision (Lead), Writing - original draft (Equal), Writing - review & editing (Equal))

## Data Availability

All data produced in the present study are available upon reasonable request to the authors

## Acknowledgments

The authors wish to acknowledge this land on which the University of Toronto operates. For thousands of years, it has been the traditional land of the Huron-Wendat, the Seneca, and the Mississaugas of the Credit. Today, this meeting place is still the home to many Indigenous people from across Turtle Island and we are grateful to have the opportunity to work on this land.

## List of abbreviations

DALY: disability-adjusted life years
ESHRE: European Society of Human Reproduction and Embryology
SOGC: Society of Obstetricians and Gynaecologists Canada
ACOG: American College of Obstetricians and Gynecologists
US: ultrasonography
MRI: magnetic resonance imaging
TNFα: tumor necrosis factor-alpha
IL: interleukin
MCP-1: monocyte chemotactic protein-1
CA125: cancer antigen 125
TVUS: transvaginal ultrasound
SPG: saline-infusion sonoPODography
TVS-C: TVUS-guided culdocentesis
Simoa: single molecule array
ELISA: enzyme-linked immunosorbent assays
VEGF: vascular endothelial growth factor
BMI: body mass index
SE: superficial endometriosis
DE: deep endometriosis
OE: ovarian endometrioma
SβG: streptavidin-β-galactosidase
RGP: resorufin-β-D-galactopyranoside
AEB: Average Enzyme per Bead
LOD: limit of detection
SD: standard deviation
IQR: interquartile range
ICAM-1: intercellular adhesion molecule-1
GRO-α: growth-related oncogene-alpha.

