## Supplemental Document for "Quantification of Protein Biomarkers in SonoPODography Fluid Using Single Molecule Arrays for Endometriosis"

**Supplemental Information**

Kezia E. Suryoraharjo^a,b^, Shay M. Freger^c^, Sophia M. Alonzi^b^, Anthony Makwanda^b^, Mathew Leonardi^c^, Alana F. Ogata^a,b,^*

^a^Department of Chemistry, University of Toronto, Toronto, ON, Canada. ^b^Department of Chemical and Physical Sciences, University of Toronto Mississauga, Mississauga, ON, Canada. ^c^Department of Obstetrics and Gynecology, McMaster University, Hamilton, ON, Canada.

**Supplemental Methods**

*Study Participants (Patients without endometriosis)*

Participants were prospectively recruited from McMaster University Medical Centre and West Lincoln Memorial Hospital, a tertiary, university-affiliated gynecologic surgical center with clinical expertise in endometriosis, chronic pelvic pain, and general gynecologic concerns. Premenopausal individuals aged 18–55 years who were scheduled to undergo laparoscopic surgery for any benign indication were eligible for inclusion. Surgical indications included, but were not limited to, hysterectomy (with or without salpingo-oophorectomy), myomectomy, and fertility-related procedures. Exclusion criteria comprised known or suspected malignancy, current pregnancy, and any contraindications to SPG and/or TVS-C. These included an obliterated rectouterine pouch, history of hysterectomy or bilateral salpingectomy, non-patent fallopian tubes (as assessed by preoperative imaging or surgical history), or extensive anatomic distortion unrelated to endometriosis (e.g., multifibroid uterus with obliteration of normal pelvic landmarks). Additional exclusion criteria were intraoperative findings precluding safe instillation or aspiration of the posterior rectouterine pouch, such as dense filmy adhesions or an inaccessible vaginal fornix. Participants from McMaster University Medical Centre who showed no visual evidence endometriosis and undergoing surgery for unrelated for unrelated benign indications were classified as controls.

Demographic and clinical information was collected at the time of enrollment using standardized case report forms and validated patient-reported questionnaires. Collected data included age, body mass index (BMI), menstrual history, pelvic pain symptoms, use of hormonal therapies, and history of prior surgical diagnosis of endometriosis. This study was approved by the institutional review board of each participating institution (UTM#44938/2023-04-10/2026-12-31, HiREB#14503/2023-03-10/2026-03-20) and adhered to the principles of the Declaration of Helsinki.

*SPG Fluid Sample Collection*

Using a TVUS probe fitted with a needle guide, a sterile echogenic 18-gauge oocyte retrieval needle was introduced through the posterior vaginal fornix into the fluid-filled pouch under direct visualization. Between 2 and 30 mL of SPG fluid was aspirated, depending on individual yield. These procedures were performed exclusively by a minimally invasive gynecologic surgeon-sonologist with advanced training in ultrasound. In the control group, composed of individuals without suspected endometriosis, SPG was similarly performed intraoperatively for sonographic assessment. SPG was followed by laparoscopic aspiration of the instilled fluid to allow comparison of biomarker profiles across disease states. In these participants, and in any endometriosis cases where TVS-C yielded insufficient fluid volume, aspiration was instead performed laparoscopically under direct visualization. Surgical aspiration was always conducted before any tissue excision or electrocautery to minimize contamination with blood, peritoneal debris, or inflammatory byproducts.

*Simoa Assays*

Bead Conjugation Buffer (Quanterix) was added to the filter to make a total volume of 500 µL. The filter device was centrifuged at 14,000 x g for 5 minutes. The flowthrough was then discarded, and the process was repeated twice. To recover the buffer-exchanged capture antibodies, the filter was inverted and placed inside a new Amicon microcentrifuge tube and centrifuged at 1,000 x g for 2 minutes. The filter was rinsed with 50 µL of Bead Conjugation Buffer and centrifuged at 1,000 x g for 2 minutes. The concentration of the capture antibodies was measured using a NanoDrop One/OneC (Thermofisher). The capture antibodies were diluted to 0.2 mg/mL in Bead Conjugation Buffer and stored on ice until ready to use. Carboxylated paramagnetic beads from the Simoa Homebrew Assay Starter Kit (Quanterix) were transferred into a microcentrifuge tube and washed with Bead Wash Buffer (Quanterix) three times to make a final concentration of 1.4 x 10^9^ beads/mL. The beads were washed with Bead Conjugation Buffer twice then resuspended in ice-cold Bead Conjugation Buffer. 1-ethyl-3-(3-dimethylaminopropyl) carbodiimide hydrochloride (EDC) (Thermofisher) reconstituted in ice-cold Bead Conjugation Buffer was added to the bead suspension to make a final concentration of 0.3 mg/mL. The beads were placed on a rotator for 30 minutes. The activated beads were then washed with Bead Conjugation Buffer once, followed by the addition of the buffer-exchanged capture antibodies and rotating for 120 minutes. The antibody-conjugated beads were washed with Bead Wash Buffer twice, resuspended in Bead Blocking Buffer (Quanterix), then placed on a rotator for 45 minutes. The blocked antibody-conjugated beads were washed in Bead Wash Buffer, followed by washing with Bead Diluent (Quanterix) and final resuspension in Bead Diluent.

Detector antibodies (VEGF cat# BAD293, R&D Systems; CA125 cat# 5609-05, R&D Systems) were biotinylated according to the manufacturer’s instructions. The detector antibodies were first reconstituted and added to a 50 kDa MWCO Amicon filter. Biotinylation Reaction Buffer (Quanterix) was added to the filter to make a total volume of 500 µL. The filter device was centrifuged at 14,000 x g for 5 minutes. The flowthrough was then discarded, and the process was repeated twice. To recover the buffer-exchanged detector antibodies, the filter was inverted and inserted into a new Amicon microcentrifuge tube and centrifuged at 1,000 x g for 2 minutes. The filter was rinsed with 50 µL of Biotinylation Reaction Buffer and centrifuged at 1,000 x g for 2 minutes. The concentration of the detector antibodies was measured using a NanoDrop One/OneC. EZ-Link NHS-PEG4-Biotin (Thermofisher) was added to the detector antibodies in 40x molar excess at a detector antibody/biotin volumetric ratio of 97.5/2.5, then incubated for 30 minutes. The biotinylated detector antibodies were then purified by adding the antibodies to a 50 kDa MWCO Amicon filter. Biotinylation Reaction Buffer was added to the filter to make a total volume of 500 µL. The filter device was centrifuged at 14,000 x g for 5 minutes. The flowthrough was then discarded, and the process was repeated four times. To recover the purified biotinylated detector antibodies, the filter was inverted and inserted into a new Amicon microcentrifuge tube and centrifuged at 1,000 x g for 2 minutes. The filter was rinsed with 50 µL of Biotinylation Reaction Buffer and centrifuged at 1,000 x g for 2 minutes.

*CA125 ELISA*

CA125 capture antibody stock (cat# DY5609-05, R&D Systems) was diluted to a working concentration of 4 µg/mL. 100 µL of working solution was added to each well. The plate was sealed and incubated overnight at room temperature (22˚C). The coated plate was washed with 1X Wash Buffer (cat# DY008B, R&D Systems) three times using a squirt bottle. Wash steps were performed in triplicate after each incubation, and all incubations were performed at room temperature. 300 µL of Reagent Diluent (i.e., blocking solution) (cat# DY008B, R&D Systems) was added to each coated well and incubated for over 1 hour at room temperature. Eight calibrators were prepared by performing serial dilutions of the recombinant protein stock (cat# DY5609-05, R&D Systems) in Reagent Diluent, along with high and low controls. 100 µL of sample was added to each well for 2 hours of incubation, followed by 100 µL of 0.1 µg/mL detector antibody for an additional 2 hours. All samples were plated in duplicates in two separate wells. 100 µL of 1X Streptavidin-HRP (cat# DY5609-05, R&D Systems) was added to each well, then the plate was covered from light for a 20-minute incubation. 100 µL of TMB ELISA Substrate (cat# DY008B, R&D Systems) was added to each well and incubated for 20 minutes, followed immediately by addition of 50 µL Stop Solution (cat# DY008B, R&D Systems) without any wash steps. Optical density was measured at 450 nm with a correction wavelength of 540 nm using a plate reader (BioTek Synergy LX, Agilent). Optical density values fit against CA125 concentrations using a 4-parameter logistic curve were used to generate calibration curves. The calibration curves were used to determine the concentrations of the samples.

*ELISA Assay Validation*

Frozen SPG samples (-80˚C) were thawed on ice. Samples from five patients were combined to constitute the pooled samples. The samples were centrifuged at 3,000 x g for 15 minutes at 4˚C, and the supernatant was collected. For dilution linearity, recombinant protein of each biomarker was spiked into the pooled sample, followed by ELISA measurements in samples serially diluted with Reagent Diluent (CA125 ELISA, cat# DY008B, R&D Systems).

Spike-and-recovery experiments were conducted on pooled samples as well as three individual samples. Percent recoveries were examined in the samples with spiked recombinant protein. For each biomarker, three concentrations were chosen at low, mid, and high-range concentrations at each dilution factor selected. The percent recoveries were calculated by subtracting the mean measured concentration of the samples without the spiked protein from the mean measured concentration of the spiked samples, then dividing by the concentration of recombinant protein spiked into the samples and multiplying by 100%.

*ELISA Biomarker Quantification*

Frozen SPG samples (-80˚C) were thawed on ice. The samples were centrifuged at 3,000 x g for 15 minutes at 4˚C, and the supernatant was collected. For quantification using ELISA assays, samples were diluted 8x in Reagent Diluent (R&D Systems). Biomarker concentrations of samples were determined using calibration curves generated on the same day of quantification.

**Supplemental Table 1**. Analytical parameters of TNF$\alpha$ Simoa calibration curves (*N* = 9).

| **LOD (pg/mL)** | **Blank signal (AEB)** | **SD of first non-blank calibrator (AEB)** | **4PL fit coefficient A** | **4PL fit coefficient B** | **4PL fit coefficient C** | **4PL fit coefficient D** |
| --- | --- | --- | --- | --- | --- | --- |
| 0.010 | 0.0062 | 0.0004 | 0.0062 | 1.034 | 4305 | 952 |
| 0.028 | 0.006 | 0.002 | 0.0057 | 1.009 | 14387 | 2784 |
| 0.014 | 0.0060 | 0.0009 | 0.0061 | 1.003 | 2210066 | 429830 |
| 0.008 | 0.0054 | 0.0005 | 0.0055 | 0.992 | 2958772 | 451080 |
| 0.003 | 0.0063 | 0.0002 | 0.0062 | 1.004 | 1939 | 374 |
| 0.012 | 0.0063 | 0.0007 | 0.0063 | 0.997 | 76206135 | 12604206 |
| 0.003 | 0.0074 | 0.0002 | 0.0074 | 1.013 | 28045 | 5196 |
| 0.006 | 0.0062 | 0.0004 | 0.0061 | 0.987 | 2183 | 370 |
| 0.003 | 0.0050 | 0.0002 | 0.0050 | 1.003 | 7957378 | 1530616 |

**Supplemental Table 2**. Analytical parameters of IL-1$\beta$ Simoa calibration curves (*N* = 9).

| **LOD (pg/mL)** | **Blank signal (AEB)** | **SD of first non-blank calibrator (AEB)** | **4PL fit coefficient A** | **4PL fit coefficient B** | **4PL fit coefficient C** | **4PL fit coefficient D** |
| --- | --- | --- | --- | --- | --- | --- |
| 0.02 | 0.008 | 0.007 | 0.0087 | 0.947 | 164 | 134 |
| 0.003 | 0.008 | 0.002 | 0.0082 | 0.948 | 205 | 193 |
| 0.008 | 0.008 | 0.004 | 0.0086 | 0.980 | 42.8 | 49.2 |
| 0.006 | 0.009 | 0.003 | 0.0084 | 0.961 | 111 | 99.7 |
| 0.005 | 0.008 | 0.002 | 0.0083 | 1.008 | 45.0 | 57.0 |
| 0.05 | 0.05 | 0.005 | 0.0250 | 1.131 | 13.3 | 22.7 |
| 0.00008 | 0.008 | 0.0002 | 0.0086 | 0.924 | 151 | 136 |
| 0.001 | 0.02 | 0.0004 | 0.0239 | 0.991 | 45.4 | 55.6 |
| 0.002 | 0.009 | 0.001 | 0.0089 | 0.943 | 141 | 108 |

**Supplemental Table 3**. Analytical parameters of VEGF Simoa calibration curves (*N* = 12).

| **LOD (pg/mL)** | **Blank signal (AEB)** | **SD of first non-blank calibrator (AEB)** | **4PL fit coefficient A** | **4PL fit coefficient B** | **4PL fit coefficient C** | **4PL fit coefficient D** |
| --- | --- | --- | --- | --- | --- | --- |
| 3.5 | 0.04 | 0.02 | 0.0416 | 1.049 | 1796 | 37.5 |
| 0.7 | 0.083 | 0.005 | 0.0854 | 1.018 | 2968 | 57.3 |
| 1.4 | 0.155 | 0.008 | 0.1549 | 1.004 | 3860 | 68.4 |
| 0.05 | 0.1705 | 0.0007 | 0.1762 | 0.928 | 2976 | 59.8 |
| 1.4 | 0.115 | 0.008 | 0.1161 | 1.059 | 1379 | 33.7 |
| 0.05 | 0.101 | 0.001 | 0.1027 | 0.981 | 9222 | 165 |
| 1.1 | 0.153 | 0.009 | 0.1574 | 1.053 | 1266 | 37.7 |
| 1.3 | 0.11112 | 0.00006 | 0.1171 | 0.987 | 3182 | 64.1 |
| 0.4 | 0.130 | 0.004 | 0.1325 | 1.034 | 1875 | 54.3 |
| 0.2 | 0.125 | 0.002 | 0.1269 | 1.042 | 1964 | 53.3 |
| 0.1 | 0.068 | 0.001 | 0.0686 | 1.043 | 3262 | 95.5 |
| 0.5 | 0.039 | 0.002 | 0.0390 | 1.050 | 2596 | 49.1 |

**Supplemental Table 4**. Analytical parameters of CA125 Simoa calibration curves (*N* = 15).

| **LOD (pg/mL)** | **Blank signal (AEB)** | **SD of first non-blank calibrator (AEB)** | **4PL fit coefficient A** | **4PL fit coefficient B** | **4PL fit coefficient C** | **4PL fit coefficient D** |
| --- | --- | --- | --- | --- | --- | --- |
| 0.01 | 0.00736 | 0.00001 | 0.0074 | 1.031 | 689 | 3.19 |
| 0.2 | 0.0069 | 0.0003 | 0.0070 | 0.971 | 1405 | 4.78 |
| 0.2 | 0.0062 | 0.0003 | 0.0063 | 0.967 | 1610 | 5.79 |
| 0.1 | 0.0120 | 0.0007 | 0.0119 | 1.000 | 682 | 3.21 |
| 0.1 | 0.0099 | 0.0003 | 0.0101 | 0.965 | 9674510 | 25326 |
| 1.0 | 0.010 | 0.001 | 0.0102 | 0.987 | 71117155 | 226011 |
| 0.08 | 0.00962 | 0.00008 | 0.0094 | 0.949 | 1207370 | 2942 |
| 0.4 | 0.0100 | 0.0007 | 0.0097 | 0.980 | 789 | 3.57 |
| 0.02 | 0.00735 | 0.00006 | 0.0077 | 0.937 | 769690021 | 1283018 |
| 0.4 | 0.009 | 0.001 | 0.0088 | 0.924 | 502612 | 1214 |
| 0.001 | 0.00971 | 0.00002 | 0.0098 | 0.946 | 708193861 | 1392329 |
| 0.06 | 0.00966 | 0.00005 | 0.0094 | 0.943 | 10030384137 | 15802523 |
| 0.3 | 0.0085 | 0.0007 | 0.0084 | 0.926 | 458270 | 1179 |
| 0.8 | 0.016 | 0.002 | 0.0159 | 0.989 | 8030276 | 49646 |
| 0.2 | 0.0155 | 0.0009 | 0.0159 | 0.986 | 62215598 | 433669 |

**Supplemental Table 5**. Demographic characteristics of patients without endometriosis.

| Characteristic | Patients without Endometriosis  (*N* = 14) |
| --- | --- |
| Age (years), mean ± SD | 40 ± 8 |
| Sex  Assigned female at birth, n (%) | 14 (100%) |
| Weight (kg), mean ± SD | 80 ± 20 |
| Height (m), mean ± SD | 1.64 ± 0.06 |
| Body-mass index (kg/m^2^), mean ± SD | 30 ± 6 |
| Gravidity, median (IQR) | 1.5 (0-3) |
| Parity, median (IQR) | 0.5 (0-2) |
| Saline volume introduced (mL), mean ± SD | 81 ± 5 |
| Estimated initial peritoneal fluid (mL), mean ± SD | 2 ± 3 (*N* = 7)* |

*Pre-operative imaging was not performed on patients without endometriosis from West Lincoln Memorial Hospital, so these volumes will remain unknown.

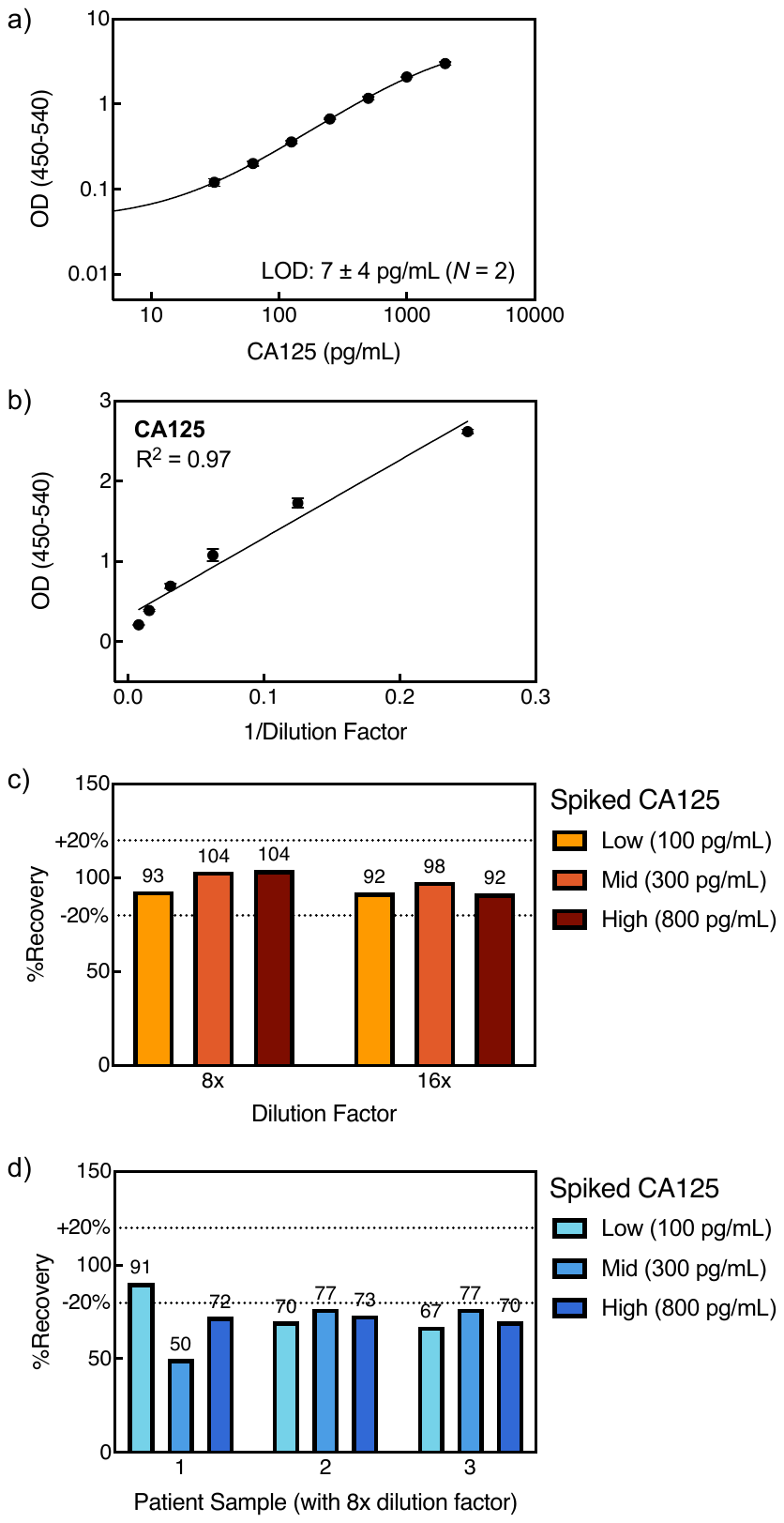

**Supplemental Figure 1**. a) CA125 ELISA calibration curve (*N* = 2). The plotted calibration curve is a representative example from *N* independently generated calibration curves. Error bars represent the SD at each concentration across all *N* replicates. LOD of the assay represent the mean of *N* independently generated LODs. b) CA125 ELISA assay response over 4x to 128x dilution factors. Data are the average of two replicates with error bars representing standard deviations. c) Percent recoveries of CA125 Simoa assay in pooled patient SPG fluid samples. d) Percent recoveries of CA125 Simoa assay in three individual patient SPG fluid samples.

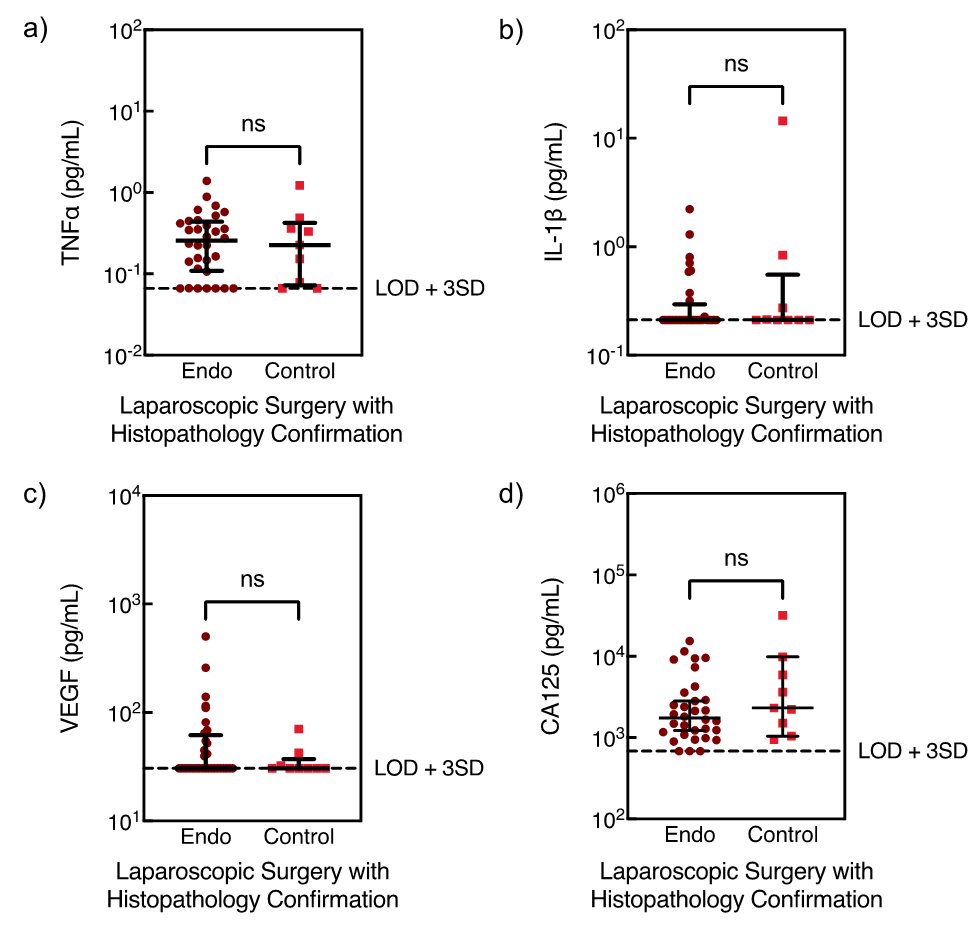

**Supplemental Figure 2.** Comparison of a) TNF$\alpha$, b) IL-1$\beta$, c) VEGF, d) CA125 concentrations in SPG fluid samples from patients with and without endometriosis. Diagnostic classification was determined by laparoscopic surgery with histopathology confirmation. Data are the average of two replicates. Lines indicate median with IQR. Dashed lines represent LOD + 3SD. Samples with hemolysis were excluded.

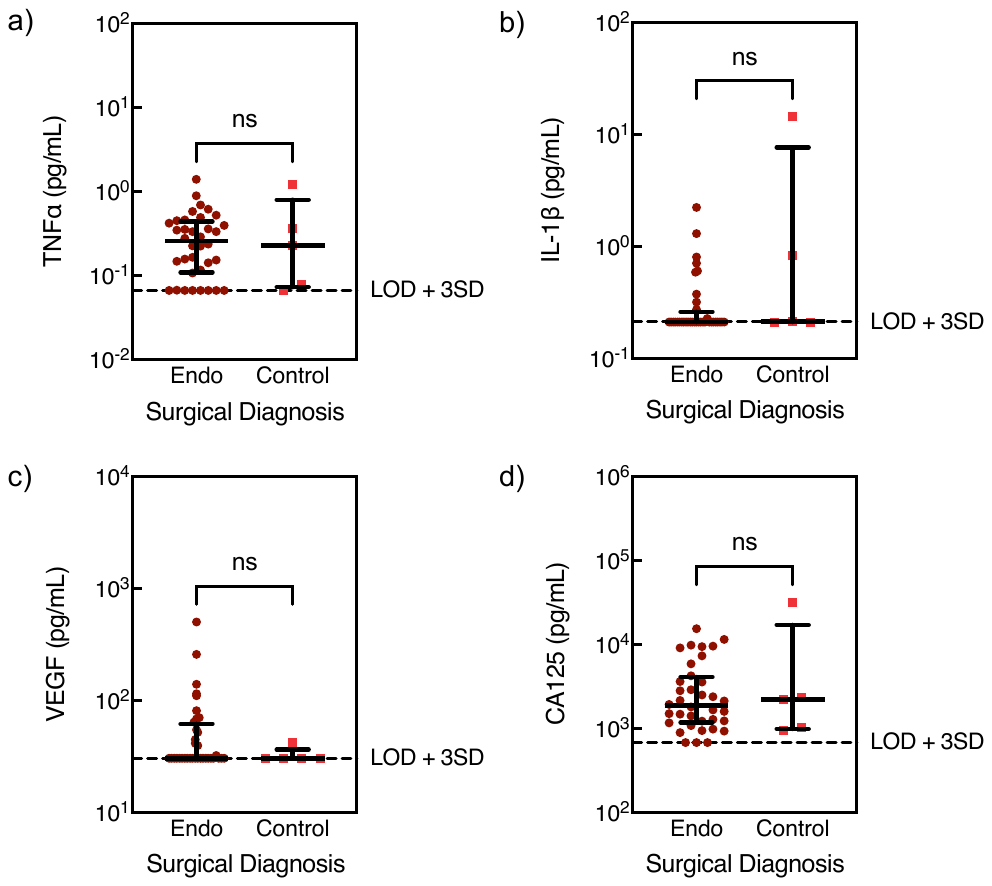

**Supplemental Figure 3.** Comparison of a) TNF$\alpha$, b) IL-1$\beta$, c) VEGF, d) CA125 concentrations in SPG fluid samples from patients with and without endometriosis. Diagnostic classification was determined by laparoscopic surgery. Data are the average of two replicates. Lines indicate median with IQR. Dashed lines represent LOD + 3SD. Samples with hemolysis were excluded.

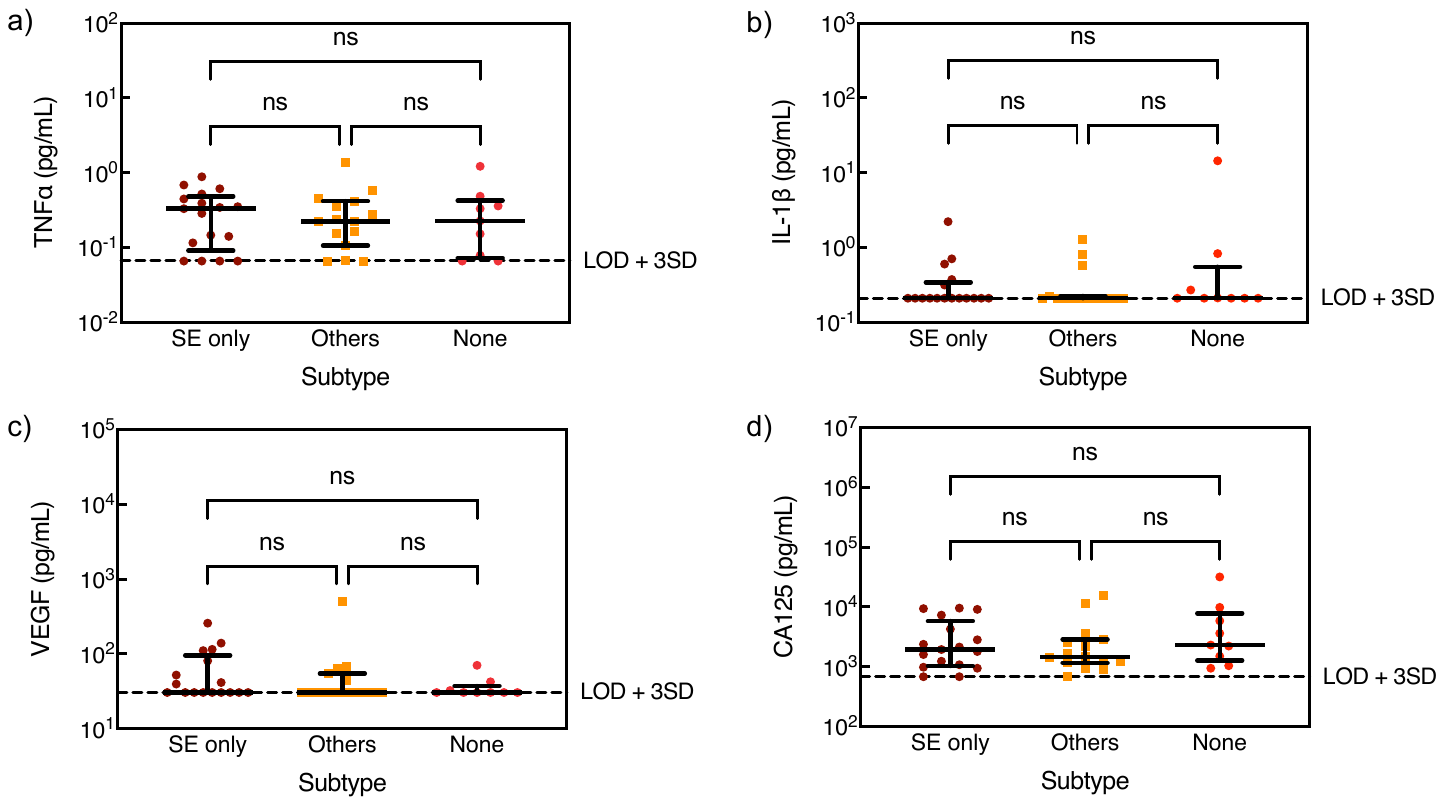

**Supplemental Figure 4**. Comparison of a) TNF$\alpha$, b) IL-1$\beta$, c) VEGF, d) CA125 concentrations in SPG fluid samples from patients with SE only, SE with other subtypes or other subtypes only (Others), and without endometriosis. Diagnostic classification was determined by laparoscopic surgery with histopathology confirmation. Identification of endometriosis subtype was done by laparoscopic surgery. Data represent the average of two replicates. Lines represent median and IQR. Dashed lines represent LOD + 3SD.

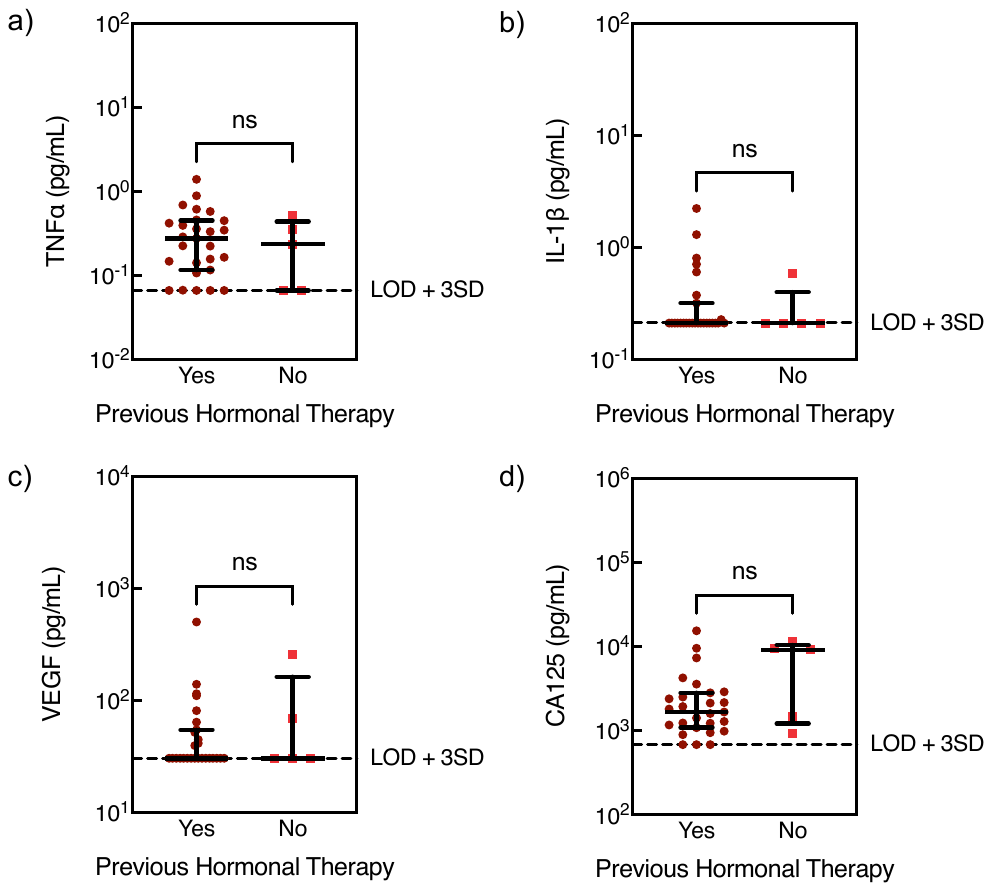

**Supplemental Figure 5**. Comparison of a) TNF$\alpha$, b) IL-1$\beta$, c) VEGF, d) CA125 concentrations in SPG fluid samples from endometriosis patients with and without previous hormonal therapy. Data are the average of two replicates. Lines indicate median with IQR. Dashed lines represent LOD + 3SD. Samples with hemolysis were excluded.

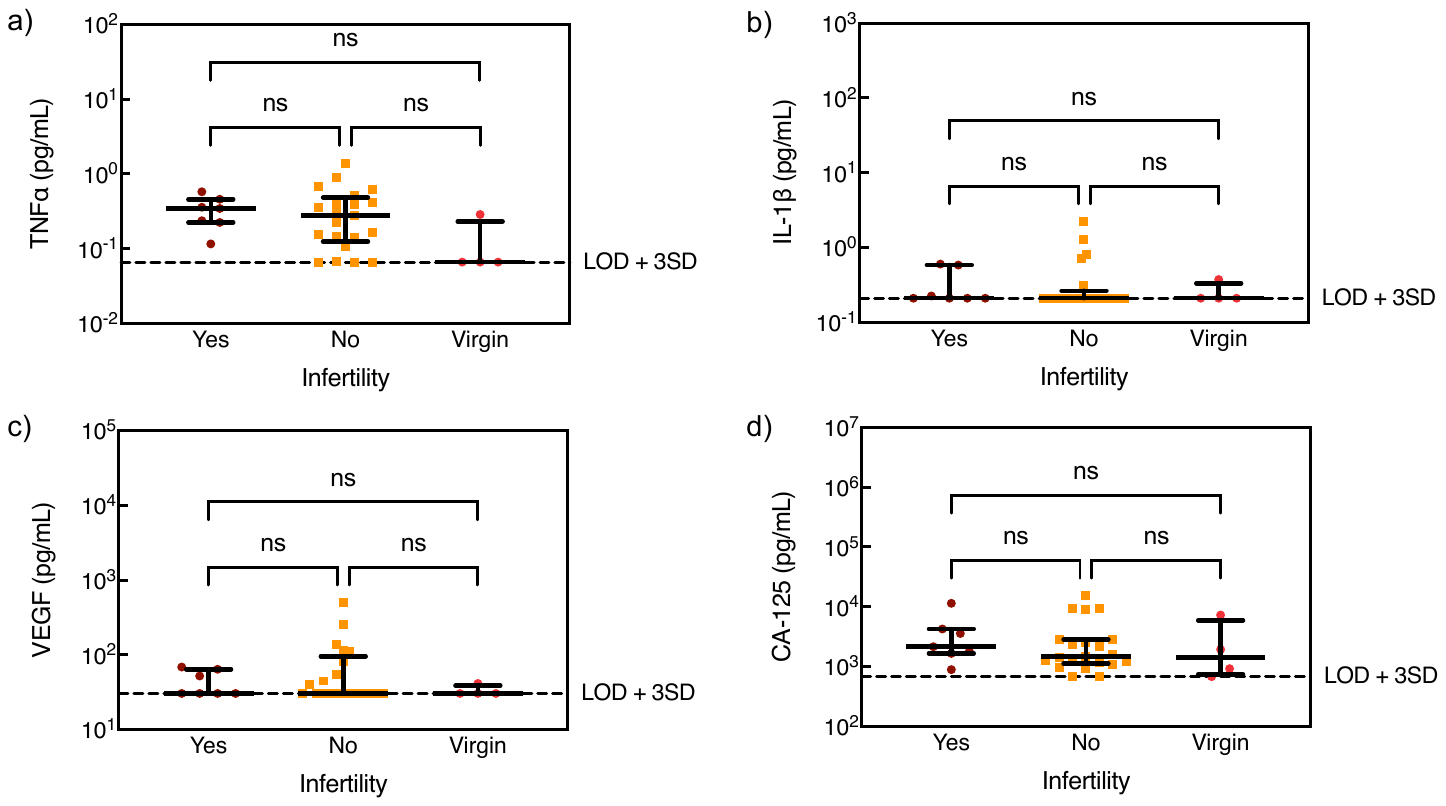

**Supplemental Figure 6**. Comparison of a) TNF$\alpha$, b) IL-1$\beta$, c) VEGF, d) CA125 concentrations in SPG fluid samples from endometriosis patients with infertility, without infertility, and virgin patients. Data are the average of two replicates. Lines indicate median with IQR. Dashed lines represent LOD + 3SD. Samples with hemolysis were excluded.

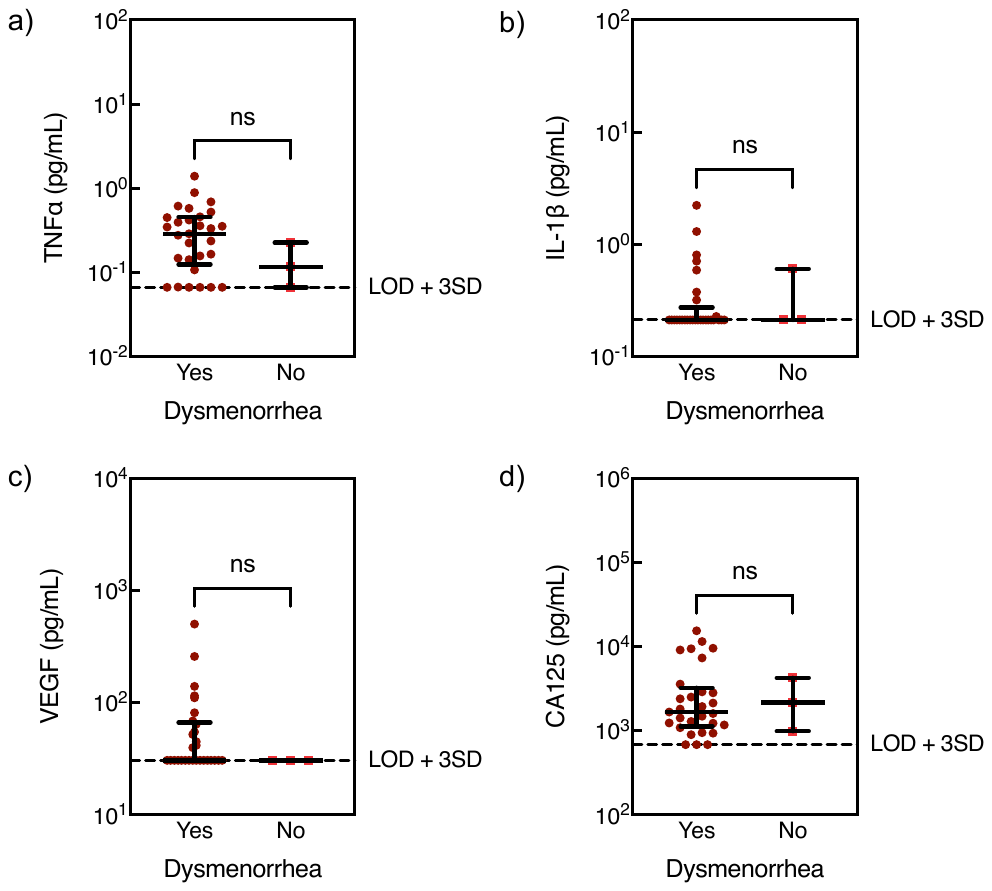

**Supplemental Figure 7**. Comparison of a) TNF$\alpha$, b) IL-1$\beta$, c) VEGF, d) CA125 concentrations in SPG fluid samples from endometriosis patients with and without dysmenorrhea. Data are the average of two replicates. Lines indicate median with IQR. Dashed lines represent LOD + 3SD. Samples with hemolysis were excluded.

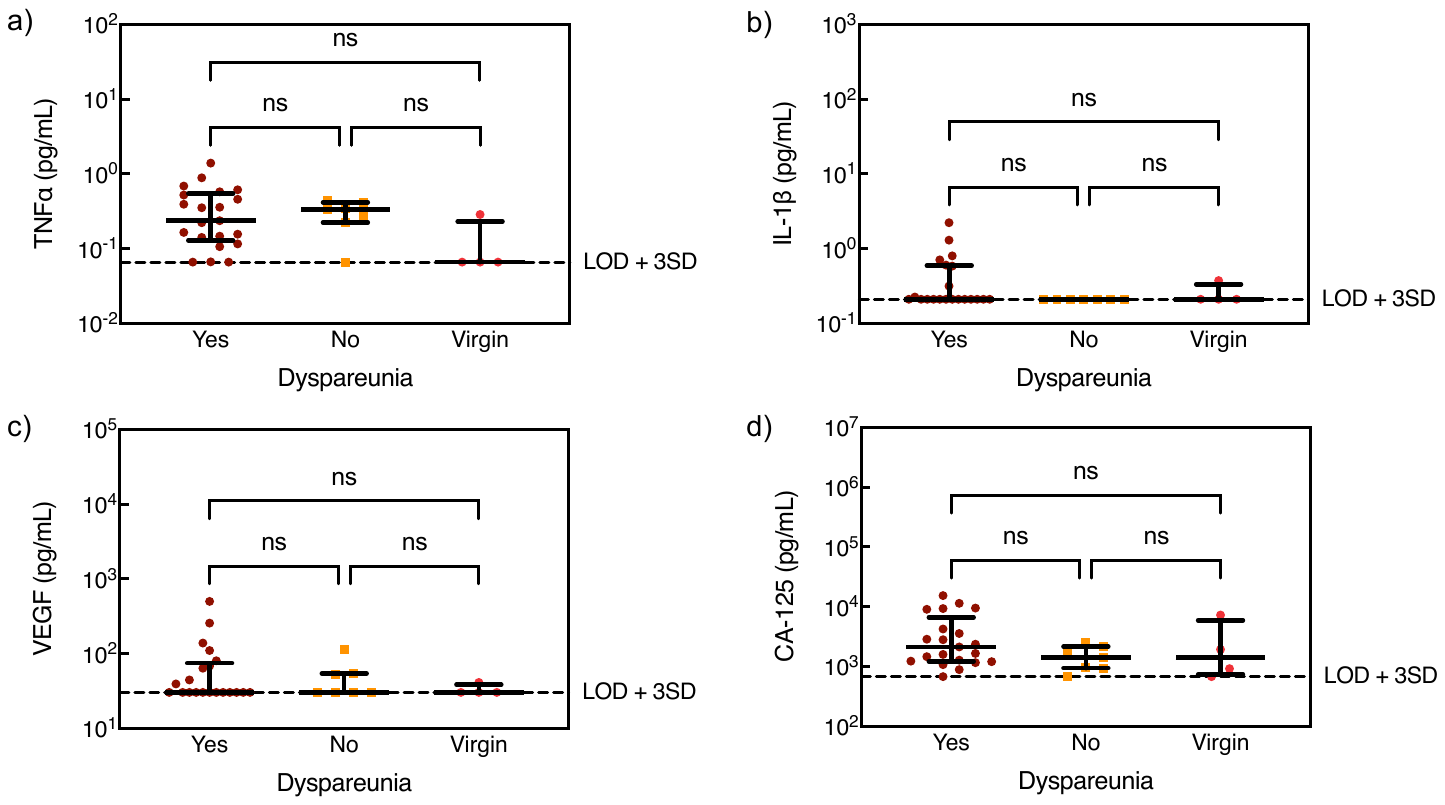

**Supplemental Figure 8**. Comparison of a) TNF$\alpha$, b) IL-1$\beta$, c) VEGF, d) CA125 concentrations in SPG fluid samples from endometriosis patients with dyspareunia, without dyspareunia, and virgin patients. Data are the average of two replicates. Lines indicate median with IQR. Dashed lines represent LOD + 3SD. Samples with hemolysis were excluded.

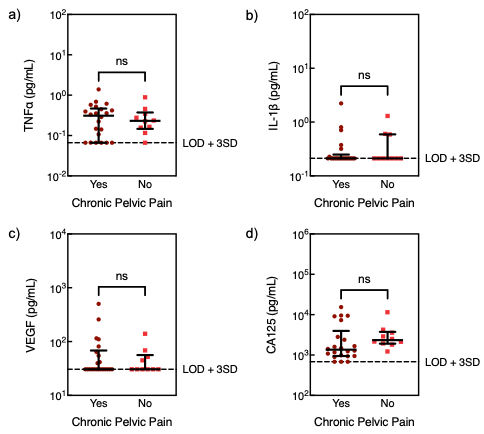

**Supplemental Figure 9**. Comparison of a) TNF$\alpha$, b) IL-1$\beta$, c) VEGF, d) CA125 concentrations in SPG fluid samples from endometriosis patients with and without chronic pelvic pain. Data are the average of two replicates. Lines indicate median with IQR. Dashed lines represent LOD + 3SD. Samples with hemolysis were excluded.
